# Gut Microbiota Markers for Antipsychotics Induced Metabolic Disturbance in Drug Naïve Patients with First Episode Schizophrenia – A 24 Weeks Follow-up Study

**DOI:** 10.1101/2020.12.26.20248886

**Authors:** Xue Li, Xiuxia Yuan, Lijuan Pang, Yu Miao, Shuying Wang, Xiaoyun Zhang, Shaohua Hu, Yunpeng Wang, Ole A. Andreassen, Xueqin Song

**Affiliations:** Department of Psychiatry, the First Affiliated Hospital/Zhengzhou University, Zhengzhou, China; Biological Psychiatry International Joint Laboratory of Henan/Zhengzhou University, Zhengzhou, China; Henan Psychiatric Transformation Research Key Laboratory/Zhengzhou University, Zhengzhou, China; Department of Psychiatry, the First Affiliated Hospital, Zhejiang University School of Medicine, Hangzhou, China; Centre for Lifespan Changes in Brain and Cognition (LCBC), Department of Psychology, University of Oslo, Forskningsveien 3A, 0373 Oslo, Norway; Norwegian Center for Mental Disorders Research (NORMENT), Institute of Clinical Medicine, University of Oslo, Norway

**Author notes:** **Corresponding Authors: Xueqin Song**, M.D., PH.D., the First Affiliated Hospital/Zhengzhou University, 450000,. These authors contributed equally to this work (Xue Li, Xiuxia Yuan).

## Abstract

**Background:** While cardiometabolic adverse effects associated with antipsychotic treatment is an important clinical challenge, the underlying mechanisms are unknown. Here we investigated if changes in gut microbial composition associate with the metabolic disturbance induced by the risperidone treatment of schizophrenia.

**Methods:** Ninety-four first episode, drug naïve schizophrenia patients (SZ), and 100 healthy controls (HCs) were enrolled at baseline. Six metabolic parameters (glucose, homeostasis model assessment of insulin resistance (HOMA-IR), Total cholesterol (Total-C), Low-density lipoprotein cholesterol (LDL-C), High-density lipoprotein cholesterol (HDL-C) and triglycerides) and body mass index (BMI) were measured for all participants. Gut microbial composition (microbials) was determined by fecal samples using 16S ribosomal RNA sequencing. Both the metabolic parameters and the gut microbiota were analyzed at baseline (all participants) and after 12 and 24 weeks of risperidone treatment (patients).

**Results:** The glucose was significantly higher in SZ than HCs at baseline (p = 0.005). After 24-weeks treatment with risperidone, the levels of BMI, glucose, HOMA-IR, Total-C, LDL-C, HDL-C and triglyceride, were significant changed compared to baseline (p < 0.01). Six microbials showed significant changes in abundance after 24 weeks of risperidone treatment in SZ (p < 0.05), and four of these (*Bacteroidetes, Proteobacteria, Christensenellaceae*, and *Enterobacteriaceae)* were associated with the changes in metabolic parameters (p < 0.05). At baseline, the abundance of the microbials *Christensenellaceae* and *Enterobacteriaceae* were significantly associated with changes in triglyceride, BMI and HOMA-IR after 24-week risperidone treatment.

**Conclusions:** Changes in gut microbial composition induced by risperidone treatment may be a key pathway underlying the metabolic disturbances observed in SZ patients. While these findings warrant replication in independent samples, they provide insight into the role of microbiota in SZ treatment, which can form the basis for development of better SZ treatment strategies.

## Introduction

Schizophrenia (SZ) is a severe mental disorder caused by both genetic and environmental factors [1,2]. Epidemiological studies have reported that the prevalence of obesity, type 2 diabetes (T2D), and hypercholesterolemia in SZ patients is 3–5 times higher than in general population [3,4]. A recent study estimated the overall rate of metabolic syndrome to 32.5% in SZ patients [3], and this rate is increased during treatment [5]. Large-scale genomic studies have revealed putative common genetic bases between SZ and metabolic disorders/traits – including cardiovascular diseases, T2D, levels of blood glucose, insulin and lipoproteins and body mass index (BMI) [6,7]. However, most second-generation antipsychotics (SGAs), recommended as first-line management for SZ [8],can generate adverse metabolic side effects, including weight gain, lipid disturbance, and glucose dysregulation [4,9]. Since most epidemiological and genetic studies included both untreated and chronic SZ patients, it is not clear whether the observed co-morbidity of SZ and metabolic syndrome is due to common genetic factors or side effects of antipsychotic treatment. Importantly, it is largely unknown by which mechanisms SGAs generate the metabolic syndrome.

Gut microbiota is considered as a “metabolic organ” greatly affecting the organism’s metabolism [10]. Several gut microbials have been shown to play important roles in the development of metabolic disorders such as T2D, obesity, hyperglycemia, and dyslipidemia [11,12]. For example, high *Firmicutes* to *Bacteroidetes* (F/B) ratio [13] and high abundance of *Actinobacteria* [14] associate with obesity. The genus *Christensenellaceae* was enriched in individuals with low BMI [15,16]. In addition, the abundance of members from *Firmicutes*, specifically the *Clostridia* class, were reduced, whereas members of *Bacteroidetes, Clostridium spp*., *Desulfovibrio spp*., *Fusobacteria* and *Betaproteobacteria* class were increased in T2D patients compared with controls [17,18].

Few studies have focused on associations between gut microbial composition and the metabolic characteristics in patients with psychiatric disorders before and after treatment with SGAs [19]. Long-term exposure to risperidone in children was associated with an increased BMI and a significantly higher F/B ratio as compared with antipsychotic-naïve psychiatric patients. *Ralstonia, Clostridium* and members of the *Erysipelotrichaceae* family were correlated with weight gain in the risperidone group [20]. Our previous study demonstrated that significant changes in certain fecal bacteria was associated with metabolic changes induced by antipsychotic medication in SZ [21].

Here, we investigated the association between gut microbial compositions with glycolipid metabolic parameters before and after 24 weeks of risperidone treatment in 94 first-episode, drug naïve SZ patients. We used 16S ribosomal RNA (16S rRNA) gene sequencing-based approach to examine the gut microbiota. And, we measured glycolipid metabolic status by serum levels of glucose, homeostasis model assessment of insulin resistance (HOMA-IR), total cholesterol (Total-C), Low-density lipoprotein cholesterol (LDL-C), High-density lipoprotein cholesterol (HDL-C), and triglycerides, which are key metabolic disturbance and traits used in SZ studies [22].

## Methods and Materials

### Participants

This is an open-pilot study lasting for 24 weeks. Participants were enrolled from the Department of Psychiatry of the First Affiliated Hospital of Zhengzhou University between October 2017 and May 2019. This study was approved by the Human Ethics Committee of the First Affiliated Hospital of Zhengzhou University, China (Approval No. 2016-LW-17). All subjects were signed a written informed consent.

The inclusion criteria for patients were: 1) met the criteria for Diagnostic and Statistical Manual of Mental Disorders fourth version (DSM-IV) schizophrenia (SZ); the Structured Clinical Interview for DSM-IV (SCID) was used to make the diagnosis[23]; 2) never treated with antipsychotics before; 3) between 18 and 45 years of age. The exclusion criteria for patients included: 1) have any physical or other mental disorders or illegal drug use; 2) pregnant or lactating women; 3) significant diarrhea or constipation in the past month; 4) a significant change in the living environment or diet in the past month; And, 5) a history of using any antibiotics, probiotics, or prebiotics in the past month.

Healthy control (HC) participants were enrolled from local communities through newspaper/online advertisement. HC must fit the same inclusion and exclusion criteria as patients except the SZ diagnosis.

### Clinical assessment

Psychiatric symptoms were evaluated by Positive and Negative Syndrome Scale (PANSS) at baseline, the 12th week, and the 24th week for patients. The reduction ratio of the PANSS score (PANSS-R) was defined as (PANSS total (PANSS-T) score at baseline minus corresponding PANSS-T score at week 24)/ (PANSS-T score at baseline minus 30).

### Treatment

All patients were treated with risperidone with the dosage gradually titrated from 1mg/day to 4-6mg/day based on clinical requirement. During the follow-up, clonazepam treatment was additionally applied to 2 patients who had sleep problems, another 6 patients had an extracorporeal vertebral response after risperidone titrated to 6mg/d, and, benzyl was taken to improve the conditions. For various reasons, not all patients completed the full study. At the 12th-week, 5 patients did not come to the assessment at the hospital; 4 patients did not provide stool sample; 3 patients discontinued medicine by themselves; another 2 patients dropped out the study due to poor treatment response. For the assessment at the 24th week, 3 patients did not come to the assessment at the hospital; 4 patients did not provide stool sample;2 patients discontinued medicine by themselves;another 2 patients discontinued medicine due to economic reasons. In total, 69 SZ patients finished the complete 24-week follow-up study (**Figure 1**).

**Figure 1.**
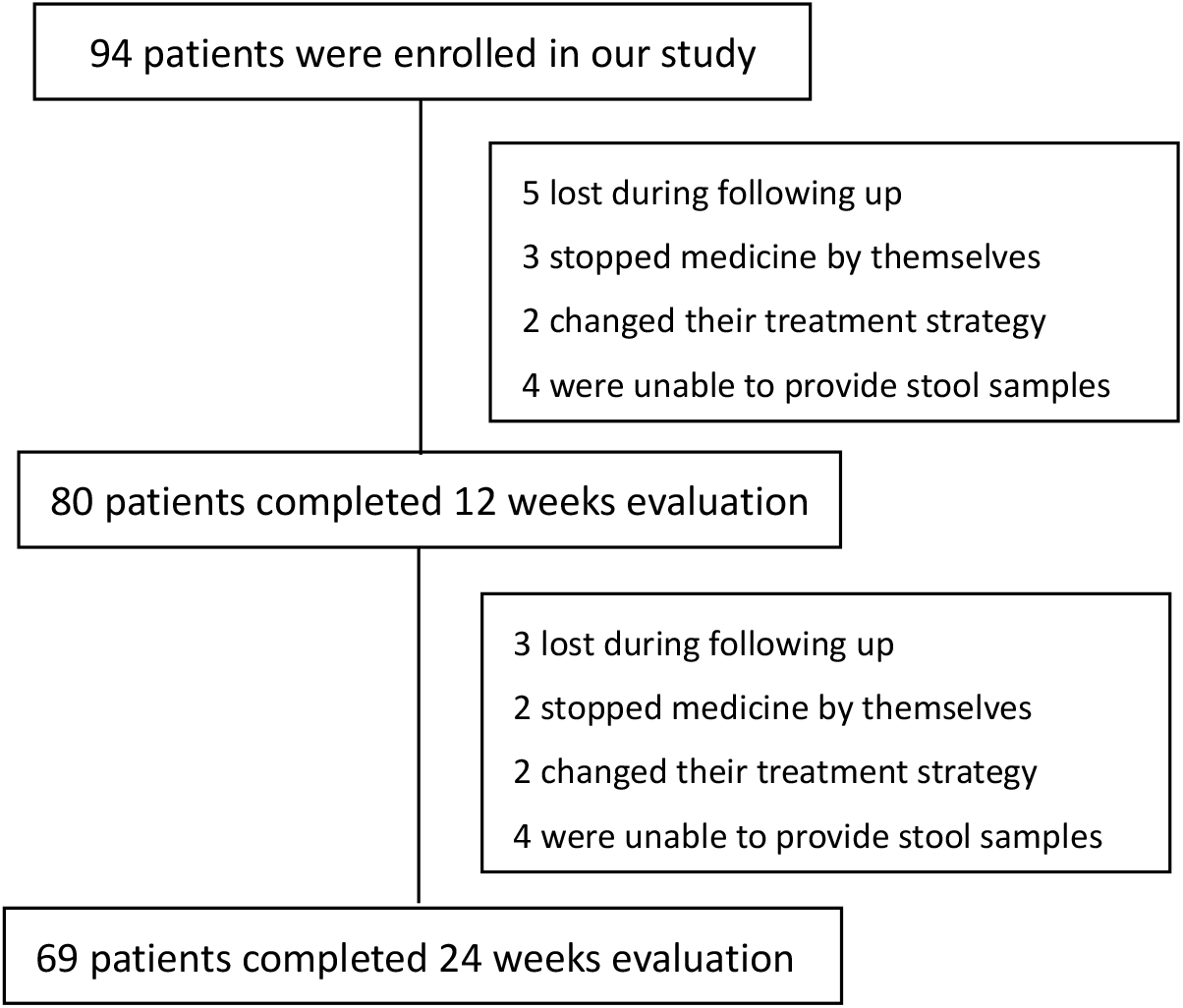
The flow chart of patients at different time points

### Biological samples collection

Venous blood (5 ml) was collected between 06:30am and 07:00am to avoid the influence of biorhythm on parameters to be measured. Serum levels of glucose,insulin, total cholesterol (Total-C), high-density lipoprotein cholesterol (HDL-C), low-density lipoprotein cholesterol (LDL-C) and triglycerides were analyzed using standard enzymatic methods and an automated analyzer (Roche Diagnostics, C8000, Germany). The homeostasis model of assessment of insulin resistance (HOMA-IR) was calculated using the following formula: [fasting serum insulin concentration (μIU/mL) × fasting glucose concentration (mmol/L) / 22.5][24]. Fresh fecal samples were collected in sterile boxes and stored at −80°C refrigerator immediately. Biological samples were collected at baseline for all participants, and at 12th week and 24th week of risperidone treatment for patients.

### DNA extraction

DNA was extracted from 0.2g of fecal sample using the Cetyltrimethylammonium Ammonium Bromide CTAB/SDS method[25]. DNA concentration and purity were monitored on 1% agarose gels. According to the concentration, DNA was diluted to 1 ng/μL using sterile water.

### Amplicon Generation and PCR products purification

The 16S rRNA gene of distinct regions (V3-V4) were amplified using specific primer with barcodes. All PCRs were carried out in 30 μL reactions with 15 μL of Phusion® High-Fidelity PCR Master Mix (New England Biolabs), and 0.2 μM of forward and reverse primers and 10 ng of template DNA. Thermal cycling consisted of initial denaturation at 98°C for 1 min, followed by 30 cycles of 98°C denaturation for 10s, annealing at 50°C for 30s, and elongation at 72°C for 30s. Lastly, 72°C elongation was applied for 5 min. After electrophoresis on 2% agarose gel, PCR products were purified using Gene JETTM Gel Extraction Kit (Thermo Scientific).

### Library preparation and sequencing

Sequencing libraries were generated using Ion Plus Fragment Library Kit 48 rxns (Thermo Scientific) following manufacturer’s recommendations. The library quality was assessed on the Qubit@ 2.0 Fluorometer (Thermo Scientific). Subsequently, library was sequenced on an Ion S5TM XL platform and 400 bp/600 bp single-end reads were generated.

### Bioinformatic analysis

High-quality clean reads were obtained by Cutadapt (V1.9.1, http://cutadapt.readthedocs.io/en/stable/) quality controlled process under specific filtering conditions. Sequence reads were compared with the reference database (Silva database, https://www.arb-silva.de/) using UCHIME algorithm (UCHIME Algorithm (http://www.drive5.com/usearch/manual/uchime_algo.html) [26] to detect and remove the chimera sequences.

Alpha Diversity The alpha diversity of the gut microbiota, indicating complexity of species diversity for a sample, was estimated by the Shannon diversity index (http://www.mothur.org/wiki/Shannon) by QIIME (Version1.7.0).

#### β Diversity

The beta diversity of the gut microbiota, estimating the differences of samples in species complexity, was calculated by QIIME software (Version 1.7.0). Principal Coordinate Analysis (PCoA) analysis was displayed by WGCNA package, stat packages and ggplot2 package in R software (Version 4.0.0).

### Statistical Analysis

SPSS version 24.0 (Armonk, NY, USA) and R software (Version 4.0.0) were used for data analysis. A p value <0.05 was used as statistical significance threshold.

### Demographics and clinical characteristics of participants and Weight Gain and microbial markers in SZ

The Kolmogorov-Smirnov (K-S) test was used to check for normality of distributions of the general (d*emographics, clinical characteristics of participants and Weight Gain and microbial markers in SZ*) abundance. For normally distributed variables, the two-sample unpaired Student’s t-test was used to test group differences; For variables that failed the K-S test, the non-parametric Mann-Whiney U test was instead used. For categorical variables, the Chi-squared test was used to test for independence between variables. The Benjamin-Hochberg (BH) was used to correct multiple testing [27].

### Clinical characteristics of SZ patients after 24-week treatment

Linear effect mixed models were used to test significant changes in clinical characteristics during 24 weeks of risperidone treatment in SZ patients. The clinical measurements were used as respond variable and follow-up duration, age, gender, duration of illness, PANSS-R were chosen as fix effects variables. The participant identifiers were used as random effect in the models. Each clinical variable was modelled separately and coefficients of follow-up durations were taken as an indicator of the changes in the variables. BH was used to correct for these testing.

### Alteration of gut microbial diversity and composition in SZ patients

The non-parametric Kruskal-Wallis test was used to evaluate the difference in microbial abundance between SZ patients and health controls (HCs). The changes in microbial markers at 12th week and 24th week of risperidone treatment in SZ were tested using similar linear effect mixed models as above but with the abundance of microbials as the respond variables.

### Microbial Markers associated with metabolic parameters at baseline (for SZ patients and HCs) and after treatment (only for SZ patients)

The Spearman rank correlations were computed for pairs of non-normally distributed variables at baseline and the changes after 24th week treatment. Partial correlation analysis was performed to explore the relationship between variables after controlling for confounding variables. Linear mixed effect models were used to measure the relationship between baseline gut microbiota and the alterations of metabolic characteristics after 24-week treatment in SZ. Here, the metabolic parameter values were used as respond variables, and microbial abundance, age, gender, duration of illness, PANSS-R were chosen as fix effects variables. The participant identifiers were used as random effect in the models. BH was used to correct for these testing.

## Results

### Demographics and clinical characteristics of participants

A total of 94 SZ patients and 100 HCs were enrolled in the study. The general clinical characteristics of the SZ patients and HCs are shown in Table 1. At baseline, there were no differences in age, gender, duration of illness, body weight, BMI, total-C, HDL-C, LDL-C and triglyceride, between the SZ patients and HCs (p > 0.05). However, serum levels of glucose were higher in SZ patients than in HCs (p = 0.005) (Table1). Among the 94 patients, 69 completed all the two time-points follow-up sampling (Figure 1). No significant difference at baseline in neither demographic nor clinical parameters was observed between the patients that dropped out of one or more follow-ups and those remained until the end of the study (p > 0.05).

**Table 1.** Demographics and clinical characteristics in patients and healthy controls at baseline

| | SZ<br>(n =94) | HC<br>(n = 100) | T/$\chi^2$ | *p* value |
| --- | --- | --- | --- | --- |
| Age(year) (mean ± SD) | 23.6±6.84 | 22.99±2.23 | 0.844 | 0.400 |
| Duration of illness (month) (mean ± SD) | 8.45±7.94 | | | |
| Gender |  |  |  |  |
| Male, n% | 71(44.6%) | 40(40%) | 0.601 | 0.438 |
| Female, n% | 88(55.4%) | 60(60%) |  |  |
| Body weight (kg) (mean ± SD) | 58.16±7.50 | 59.78±11.76 | -1.106 | 0.270 |
| BMI (kg/m<sup>2</sup>) (mean ± SD) | 20.80±1.73 | 21.19±2.74 | -1.162 | 0.247 |
| Glucose (mmol/l) (mean ± SD) | 4.60±0.98 | 4.23±0.83 | 2.813 | 0.005 |
| Total cholesterol (mmol/l) (mean ± SD) | 3.67±0.64 | 3.73±0.67 | -0.598 | 0.551 |
| Triglyceride (mmol/l) (mean ± SD) | 0.81±0.32 | 0.88±0.31 | -1.535 | 0.126 |
| HDL-C (mmol/l) (mean ± SD) | 1.32±0.28 | 1.31±0.36 | 0.227 | 0.820 |
| LDL-C (mmol/l) (mean ± SD) | 2.12±0.58 | 2.04±0.50 | 1.079 | 0.282 |

HDL-C: High-density lipoprotein cholesterol; LDL-C: Low-density lipoprotein cholesterol. Body mass index (BMI)

### Clinical characteristics of SZ patients after 24-week treatment

After controlling the effect of age, gender, duration of illness, PANSS-R, we found that levels of glucose, HOMA-IR, triglyceride, total-C, HDL-C, LDL-C and BMI changed significantly at the 12th and 24th week follow-up compared to baseline (p < 0.01). Post hoc tests revealed that glucose, HOMA-IR, triglyceride, Total-C, LDL-C and BMI increased significantly at each follow-up time point (p < 0.01) compared to baseline. HDL-C decreased significantly at 12th week and 24th week (p = 0.006, and p < 0.001, respectively) compared to baseline. (Figure 2).

**Figure 2.**
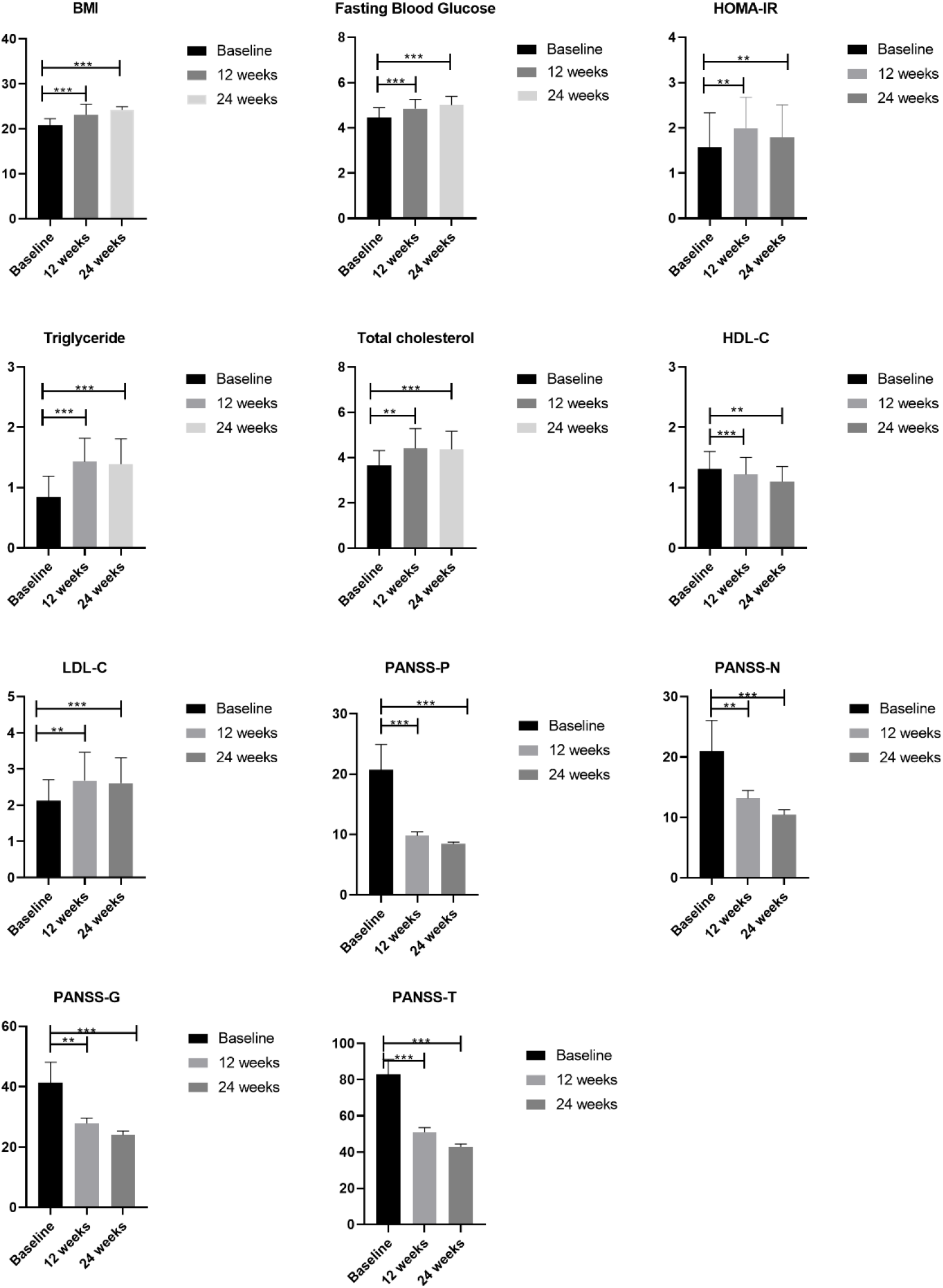
Changes of clinical characteristics of SZ patients at after 12 and 24 weeks of treatment compared to baseline. PANSS-P, PANSS-N, PANSS-G and PANSS-T were concluded in PANSS as outcome evaluation and a higher score indicates increased symptom severity. *:p<0.05,**:p<0.01,***:p<0.001

As expected, the PANSS total and subscale scores were significantly decreased at each follow-up time point during the 24 weeks risperidone treatment in patients (p < 0.01) (Figure 2).

### Gut microbial markers in SZ patients versus HC and after treatment

We did not observe significantly differences in alpha diversity, measure by Shannon diversity index, between SZ patients and HCs at baseline (p > 0.05). We found a statistically higher β diversity, calculated using Bray-Curtis distance in SZ patients than in HCs at baseline (p < 0.001) (Supplemental Figure1). At phylum level, there were lower level of *Bacteroidetes* (Z = −9.42, p < 0.001), higher levels of *Firmicutes* (Z = − 11.35, p < 0.001) and *Actinobacteria* (Z = −11.74, p < 0.001) in SZ than those in HCs at baseline. At microbiota family level, we discovered 17 families that significantly correlated with the caseness of participants based on the Wilcox test (p < 0.05). In addition, there were no significant difference in abundance of *Proteobacteria* phylum and 9 families between SZ patients and HCs (the average abundance>10^−5^,p > 0.05).

After 24-week risperidone treatment, there was no significantly differences in Shannon diversity index in the following time points (12-weeks and 24-weeks after treatment) of SZ compared to those at baseline (p > 0.05). The β diversity showed significantly differences among the following time points of SZ (p < 0.001).

Furthermore, we found that β diversity decreased significantly at 12th week and 24th week (p < 0.001) compared to baseline (Supplemental Figure1). We compared these bacteria at baseline in SZ patient with those at the two follow up time points after controlling the effect of age, gender, duration of illness, PANSS-R. At phylum level, a lower abundance of *Bacteroidetes* (t = 5.69, p < 0.001) and a higher abundance of *Proteobacteria* (t = −2.85, p = 0.006) were found after 24 weeks treatment. At family level, we discovered that there was significantly lower abundance of *Christensenellaceae* (t = 3.03, p = 0.002) and *Prevotellaceae* (t = 2.15, p = 0.033), higher abundance of *Enterobacteriaceae* (t = −2.40, p = 0.019) and *Lachnospiraceae* (t = −2.86, p = 0.005) after 24 weeks treatment.

### Gut microbial markers associated with metabolic parameters in SZ patients and HCs at baseline

We found that fecal abundance of family *Enterobacteriaceae* was positively associated with Total-C (r = 0.234, p = 0.036) and LDL-C (r = 0.234, p = 0.036) in HC. But in SZ patients, the abundance of family *Enterobacteriaceae* was positively associated with serum levels of triglyceride (r = 0.268, p = 0.018), glucose (r = 0.385, p = 0.001), and HOMA-IR (r = 0.352, p = 0.002). The abundance of family *Christensenellaceae* was negatively associated with triglyceride (r = −0.229 p = 0.034) only in SZ patients. The abundance of family *Lachnospiraceae* was positively associated with the serum levels of glucose (r = 0.236 p = 0.039), HOMA-IR (r = 0.266, p = 0.020) and negatively associated with HDL-C (r = −0.268, p = 0.018) in SZ patients. The abundance of family *Bacteroidaceae* was negatively associated with the serum levels of glucose (r = −0.250, p = 0.024) in HCs.

We found that the phylum *Bacteroidetes* (r = −0.248, p = 0.026) was negatively associated with the serum levels of glucose in HCs. The abundance of phylum *Proteobacteria* was negatively associated with HDL-C (r = −0.306, p = 0.005) in HCs. But in SZ patients, the abundance of phylum *Proteobacteria* was positively associated with the serum levels of total cholesterol (r = 0.251, p = 0.026), LDL-C (r = 0.305, p = 0.007) and triglyceride (r = 0.269, p = 0.017). Our results showed that there was a positive association between the fecal abundance of phylum *Actinobacteria* and the serum levels of total-C (r = 0.241, p = 0.030) in HCs. But in SZ, the abundance of phylum *Actinobacteria* was positively associated with serum levels of triglyceride (r = 0.413, p < 0.001) and glucose (r = 0.378, p = 0.001) (Figure 3).

**Figure 3.**
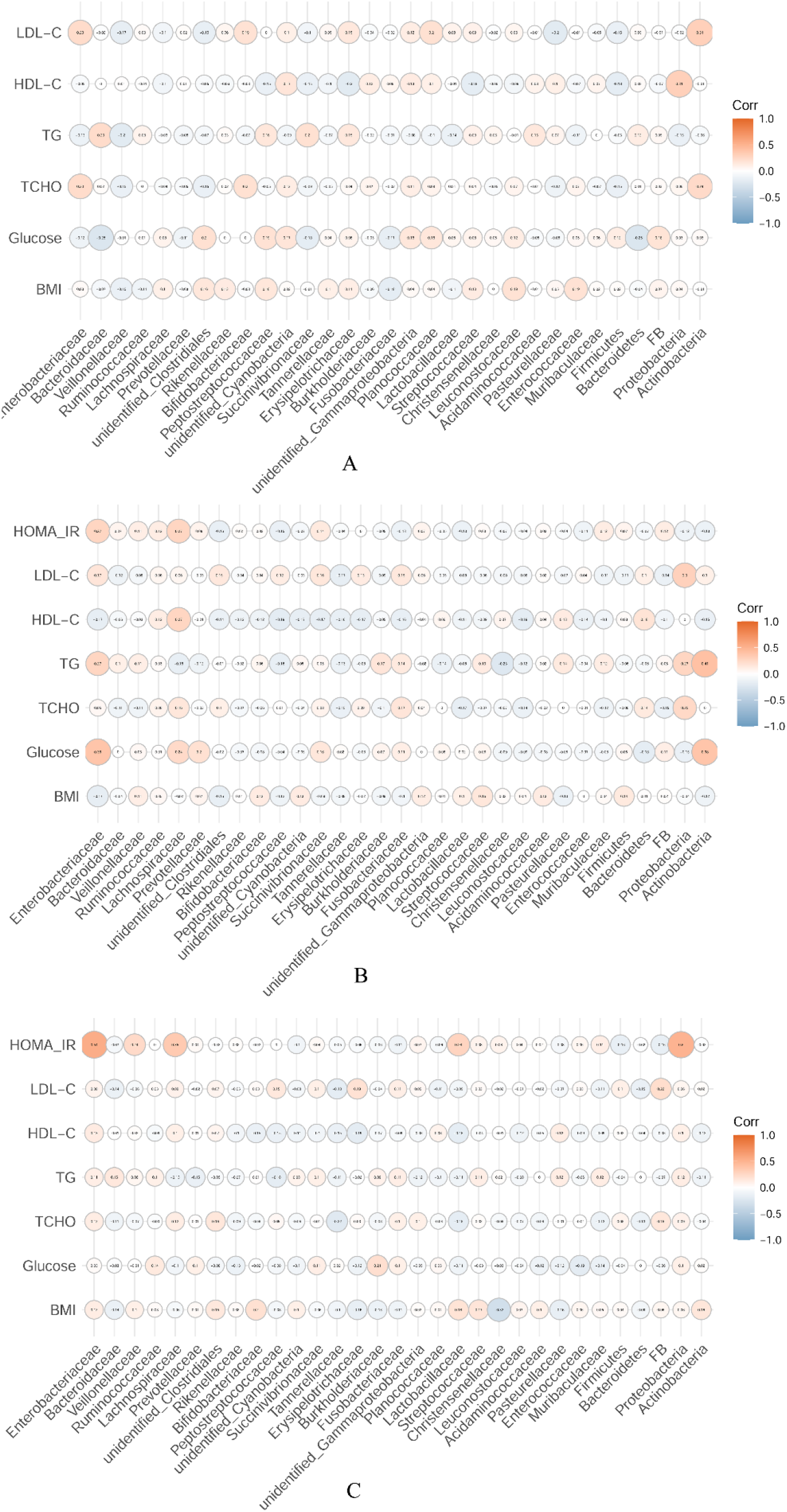
Microbial markers associated with metabolic parameters in SZ patients and HCs A: Microbial markers associated with metabolic parameters in HCs; B: Microbial markers associated with metabolic parameters in SZ patients at baseline; C: Microbial markers associated with metabolic parameters in SZ patients after 24-week treatment. The orange circle represents a positive correlation, and the blue circle represents a negative correlation. The size of the circle represents the strength of the correlation. The text on the circle is the coefficient of association.

### Changes of microbial markers associated with metabolic parameters after treatment in SZ

We investigated if there was any association between the change of gut microbiota and metabolic parameters after controlling for the effect of age, gender, duration of illness, PANSS-R in SZ patients after treatment. Our results showed that the change of family *Enterobacteriaceae* was positively associated with the change of triglyceride levels (r = 0.64, p = 0.015) and that of HOMA-IR (r = 0.61, p = 0.021) after 24-week treatment. The change of family *Christensenellaceae* was negatively associated with the change of triglyceride levels (r = −0.84, p < 0.001) and the change ratio of BMI (r = −0.83, p < 0.001) after treatment. The change of phylum *Proteobacteria* was positively associated with the change of triglyceride levels (r = 0.583, p = 0.029), HOMA-IR (r = 0.67, p = 0.008), Total-C level (r = 0.69, p = 0.007) and LDL-C (r = 0.76, p = 0.002) after treatment. The change of phylum *Bacteroidetes* was negatively associated with the change of triglyceride levels (r = −0.556, p = 0.025) after treatment. (Figure 3). Furthermore, linear mixed effect models were used to study whether baseline gut microbiota associate with the change of metabolic parameters after treatment. We found that a low fecal abundance of *Christensenellaceae* family at baseline contribute to a significant elevated of triglyceride (t = −3.46, p = 0.003) and BMI (t = −2.84, p = 0.01) after 24-week risperidone treatment. Moreover, that baseline fecal abundance of *Enterobacteriaceae* was significant associated with the change of HOMA-IR after 24-week risperidone treatment (t = 3.18, p = 0.004).

### Weight Gain and microbial markers in SZ

Based on the significant increase of BMI after 12-week treatment, SZ patients were divided into weight gain (WG) group and non-weight gain (non-WG) group using 7 % weight gain [8] at 12th week as a cutoff value. At 12th week, the WG group (N = 54) had significantly higher levels of glucose, HOMA-IR, Total-C, triglyceride, HDL-C, and LDL-C compared with the non-WG group (N = 26) (p < 0.01). There were higher abundance of *Enterobacteriaceae* (Z = −4.68, p < 0.001) and lower abundance of *Christensenellaceae* (Z = −2.76, p = 0.006) in the WG group compared with those in the non-WG group (Table 2).

**Table 2.**
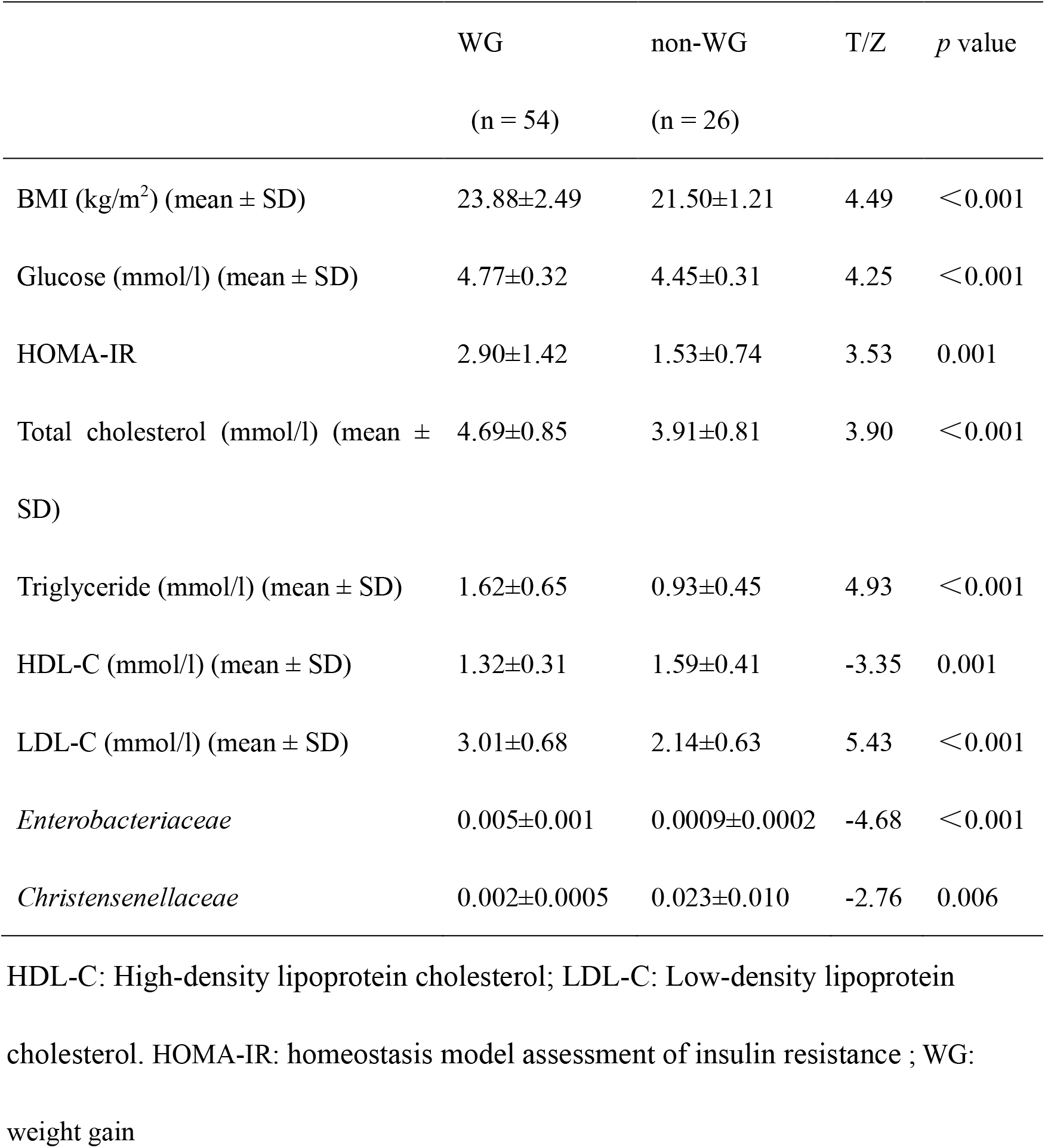
Clinical characteristics and gut microbiota at 12th week of treatment in SZ with and without weight gain

## Discussion

Here, we found indications that the use of antipsychotics modifies the ecosystem of the gut in patients with SZ. Such modifications were manifested in both diversities of the residential microbials and in specific phyla and families. Concordantly, for SZ patients, several metabolic parameters and 6 microbials significantly changed, compared to baseline, during the 24th week risperidone treatment. Among these 6 microbials, *Bacteroidetes, Proteobacteria, Christensenellaceae*, and *Enterobacteriaceae* were associated with the changes in metabolic parameters after 24 weeks of treatment for SZ patients. Further, SZ patients with weight gain showed significantly higher microbial disturbances, higher abundance of *Enterobacteriaceae* and lower abundance of *Christensenellaceae*, compared to non-weight gain SZ patients at the 12th week treatment for SZ patients. Remarkably, the changes in the abundance of two microbials, *Christensenellaceae* and *Enterobacteriaceae*, associated with changes in several metabolic parameters studied here. Together, the current findings suggest a role of gut microbiota in cardiometabolic adverse effects of antipsychotic treatment.

Consistently with animal data[28], human studies have consistently demonstrated that the *Christensenellaceae* family is increased in people with low BMI[16,29]. In a gut microbiome analysis of 1,126 twin pairs, the family *Christensenellaceae* (phylum Firmicutes) was highly heritable taxon among common taxa [15]. Our results further indicate that risperidone treatment may decrease the abundance of family *Christensenellaceae* in the gut, thereby increased the BMI for SZ patients. We also observed a negative correlation between the *Christensenellaceae* family and the levels of the triglyceride. Previous studies have shown the comorbidity between metabolic syndrome and SZ [3-5], and common genes may underlie such observed comorbidity [30]. Large-scale genomic studies have shown that there are a handful of pleiotropic genes associated with triglyceride, BMI and SZ [6]. Since large-scale genomic studies showed opposite effect directions between BMI and SZ[31], our results argue that the comorbidity between SZ and metabolic syndrome is due to changes in the environment of the gut microbiota induced by antipsychotic treatment. Most of the SGAs treatment induce a range of metabolic disturbance in SZ patients [9]. Our study design with pre-treatment baseline measure of antipsychotic-naïve patients in comparison with status after 24 weeks of treatment enabled us to identify antipsychotic-induced effects. Furthermore, we found that a low fecal abundance of family *Christensenellaceae* at baseline may contribute to a high triglyceride or BMI after 24-week risperidone treatment for SZ. Despite that these associations cannot provide for causal pathophysiology; it has been suggested that abnormal *Christensenellaceae* may influence the lipid metabolism after risperidone treatment in patients[32].

Previous study on T2D and intestinal permeability showed that enrichment in *Enterobacterial* may characterize impaired colonic permeability prior to/independently from a disruption in glucose tolerance[33]. The fecal abundance of enterobacterial ClpB gene in the microbiota correlated with BMI or obese [34]. We found that fecal abundance of *Enterobacteriaceae* was associated with triglyceride and HOMA-IR in SZ patients, which is in agreement with a previous report investigating the relationship between insulin and *Enterobacteriaceae* in infant born from an obesity mother [35]. Our results showed that baseline fecal abundance of *Enterobacteriaceae* was significantly associated with the change of HOMA-IR after risperidone treatment in patients with SZ. However, the biophysiological mechanisms by which *Enterobacteriaceae* affects glycolipid metabolism, especially in HOMA-IR and triglyceride after risperidone treatment in SZ need further investigation.

The abundance of phylum *Bacteroidetes* and *Proteobacteria* were involved in the pathophysiology of metabolic syndrome [36]. Many studied have demonstrated that *Bacteroidetes* is correlated with the change of weight and might have potential therapeutic implications in obesity [37,38]. Our results showed that the change of *Bacteroidetes* was negatively associated with the change of triglyceride after 24-week treatment in SZ patients. Previous study showed that T2D induces a gut dysbiosis, characterized by a decrease in the butyrate-producing bacteria abundance including the *Bacteroidetes* [39], which is consistent with our result that there was a negative association between *Bacteroidetes* and serum level of glucose in HCs. It dictated that the association between *Bacteroidetes* and glycolipid metabolism was found not only in healthy controls, but also in schizophrenia. Previous studies suggested that there was an intimate association between an abundance of *Proteobacteria* in the microbiota and the difficulty for the host of maintaining a balanced gut microbial community. *Proteobacteria* were demonstrated with T2D, obesity, metabolic syndrome and other metabolic diseases[40]. Our results are consistent with previous studies and showed a significant association between metabolic parameters and *Bacteroidetes, Proteobacteria* after 24 weeks of treatment with risperidone. It indicated that there may has a powerful connection between metabolic disturbance and *Bacteroidetes, Proteobacteria* no matter induced by antipsychotics or metabolic syndrome.

Several researchers have reported that *Lachnospiraceae* was associated with SZ symptom severity [41,42]. It is also believed that *Lachnospiraceae* plays a vital role in the inflammation of obesity [43]. Our results showed that *Lachnospiraceae* was positively associated with glycolipid metabolism parameters only in SZ patients at baseline. Whether the *Lachnospiraceae* is involved in the pathophysiology of SZ through the immune pathway need further investigation. However, several lines of evidence suggest a role of immune system in SZ pathology and treatment[44] (REF) and the current findings are in line with the hypothesis that the microbiota interplays with the immune system and thus contribute to cardiometabolic comorbidity in SZ.

One strength of our study is the use of drug-naïve, first-episode schizophrenia patients, which minimizes potential confounding, such as previous exposure to antipsychotics, and metabolic disturbance at baseline. Furthermore, our longitudinal study design followed patients for a period of 24 weeks. Thus, changes in the composition of gut microbiota may truly reflect the effect of metabolic disturbance after 24-week risperidone treatment rather than stochastic fluctuations. However, our results should be interpreted with limitations in mind. Our study did not assess the relative contributions of the lifestyle, especially diet habits. However, most of our participants were from Henan province, China, and had similar dietary habits for cooked wheaten food. We only focused on one common antipsychotic treatment of SZ, i.e., risperidone. Thus, whether our results can be generalized to other SGAs need further studies. Since our study has a relatively long follow-up time, some participants dropped out during follow-up. However, we performed sensitivity analysis for the drop-outs and we did not observe any distinct patterns of the drop-out group compared to the remaining samples in neither metabolic parameters nor clinical symptoms. Future studies with larger sample size and longer follow-up period are warranted to replicate our findings.

In conclusion, the present findings indicate that the metabolic side-effects of antipsychotic treatment for SZ involve the modification of the ecosystems of the gut microbiome, especially the abundance of *Christensenellaceae* and *Enterobacteriaceae*. Future development of antipsychotics should take into account their potential impact on the gut microbiota.

## Data Availability

The data of our study would be available if its needed.

## Acknowledgements

The authors thank all the patients and their families, the healthy volunteers for their participation, and the psychiatrist who helped us take clinical data and blood samples in the Department of Psychiatry, the First Affiliated Hospital of Zhengzhou University. We thank the Biological Sample Bank of The First Affiliated Hospital of Zhengzhou University for helping us store the samples. We thank the Academy of Medical Science, Zhengzhou University for providing us with a perfect experimental platform.

